# Protein-based tools for the detection and characterisation of Oropouche virus infection

**DOI:** 10.1101/2025.02.26.25322767

**Authors:** Monique K. Merchant, Juliano de Paula Souza, Sahar Abdelkarim, Shrestha Chakraborty, Tufi A. Nasser Neto, Kristel I. Gutierrez Manchay, Daniel M. de Melo Jorge, Valdinete Alves do Nascimento, Yuxian Sun, Eve R. Caroe, Lauren E.A. Eyssen, Jared S. Rudd, Isa C. Ribeiro Piauilino, Sérgio Damasceno Pinto, Matheus H. Pereira da Silva, Felipe Rocha do Nascimento, Filipe Gomes Naveca, José L. Proenca Modena, Luis L.P. daSilva, Regina M. Pinto de Figueiredo, Raymond J. Owens, Eurico Arruda, Stephen C. Graham

## Abstract

Oropouche fever is a neglected tropical viral disease endemic to Latin America and the Caribbean. Oropouche fever is caused by Oropouche virus (OROV), an orthobunyavirus with a tri-segmented (-)ssRNA genome. A recent epidemic caused by a novel reassortant has seen the first recorded deaths from OROV infection and reports of vertical transmission, affecting individuals across a dramatically expanded geographical range. Despite its importance as an emerging pathogen, research into OROV infection is hampered by paucity of available tools for serology and molecular virology. We have purified high-quality recombinant OROV nucleoprotein and the spike region of the viral surface glycoprotein Gc. We have used these antigens in indirect ELISA to detect seroconversion following experimental infection of animals, confirming their antigenic authenticity. These antigens stimulate the production of high neutralizing antibody titres in animals, highlighting their promise as immunogens for vaccination. We generated nanobodies that recognize both recombinant and infection-derived OROV antigens and developed a sandwich ELISA assay that can detect OROV antigens in human clinical serum samples with high efficiency. We have also shown that recombinant nanobodies directed against OROV Gc spike can potently neutralise infection by both historical OROV strains and the newly emerged reassortant. These protein-based reagents will accelerate OROV research and highlight the utility of protein-based tools for future OROV vaccines and point-of-care diagnostic devices.

## Background

Oropouche virus (*Orthobunyavirus oropoucheense*; OROV) is the cause of Oropouche fever, an arthropod-borne disease endemic to Latin America and the Caribbean [1,2]. First identified in Trinidad and Tobago in 1955 [3], OROV causes acute illness characterised by fever, headache, myalgia and arthralgia [4], and it can cross the blood-brain barrier to cause aseptic meningoencephalitis [5,6]. Despite causing numerous outbreaks in the Amazon basin over the last 65 years (reviewed in [7,8]), OROV remains a neglected tropical disease. The similarity of symptoms of OROV infection to those caused by co-circulating arboviruses like Dengue and Chikungunya viruses prevent the diagnosis based purely on clinical presentation and mean that the prevalence of OROV infection is likely to be underestimated [2,5,9].

Since late 2023 there has been a dramatic increase in the number of OROV cases detected across Brazil, Bolivia, Colombia, Peru and Cuba [10,11]. OROV has a tri-segmented RNA genome and this increase in cases is likely to have been driven by the emergence of a novel reassortant [12,13]. While the primary clinical presentation of infection by this new epidemic reassortant is similar to that seen in previous outbreaks [13], there have been several reports of vertical transmission in Brazil that may have caused foetal death, miscarriage or microcephaly of the newborn [14,15]. This outbreak has also seen the first two fatal OROV infections in adults, with two women in their 20s from a nonendemic region of Brazil succumbing to the infection [16]. Notably, in both fatal cases the clinical course was remarkably similar to that of severe Dengue virus infection, emphasising the need for definitive molecular diagnosis to correctly ascribe severe febrile illnesses aetiology.

Diagnosis of OROV infection can be achieved via RT-PCR analysis of serum samples collected within 5 to 7 days of symptom onset [17–19], and RT-PCR analysis can be multiplexed to also detect co-circulating viruses like Mayaro virus [20]. However, RT-PCR analysis is often unavailable in primary points of care in epidemic regions and OROV infection is not always considered, leading to significant underdiagnosis [21,22]. Multiple groups have developed in-house serological tests for OROV infection, including ELISA and immunofluorescence assays (summarised in [7]), but these have only recently become commercially available. A recent study raised monoclonal antibodies that could detect OROV infection in experimentally infected mice (immunohistochemistry) or cultured cells (immunocytochemistry), but its utility for detecting acute OROV infection in humans was not determined [23].

The dearth of detection reagents for OROV antigens impedes the development of point-of-care diagnostics tests like lateral-flow devices and slows the pace of OROV molecular virology research. In this study we generated a panel of Variable Heavy-chain domains of camelid Heavy-chain antibodies (VHHs; a.k.a. nanobodies) [24] that specifically detect the nucleoprotein and surface glycoprotein of OROV. Bivalent formulations of these nanobodies can detect OROV antigens in the serum of patients with acute OROV infection and potently neutralise OROV infection *in vitro*. These detection reagents open new avenues for the development of low-cost protein-based detection assays to detect and antiviral therapies to treat OROV infection. Furthermore, we show that the antigens used to generate the nanobodies elicit strong neutralising antibody titres in mice, highlighting their potential for use as vaccine immunogens.

## Methods

### Plasmids

The N protein (GenBank AJE24680.1) and Gc spike region (residues 482–894) of the M polyprotein (GenBank AJE24679.1) from OROV strain BeAn19991 [25], and the Gc spike region (residues 478–906) of the M polyprotein from Cristoli virus (CRIV; GenBank MN488997.1) [26], were ordered as codon-optimised synthetic genes (GeneArt). OROV N was cloned into pOPTH, derived from pOPT [27] and encoding an N-terminal His_6_ tag, or into pMWCAVI encoding an N-terminal His_6_ tag, human rhinovirus 3C protease site and biotin acceptor peptide (AviTag). The OROV and CRIV Gc spikes with N-terminal secretion signals and C-terminal His_6_ tags were cloned into the vector PB-TSW, which is derived from PB-T-PAF [28] after the insertion of a woodchuck hepatitis virus (WHV) posttranscriptional regulatory element (WPRE) at the 3′ end. The pF5A_PBase plasmid encoding the piggyBac transposase (PBase) was generated by subcloning the PBase sequence from pPBase [28] into the vector pF5A (Promega). The PB-RN plasmid carrying the reverse tetracycline transactivator inducer and neomycin resistance gene was previously described [28]. A second OROV Gc spike construct was generated with a C-terminal AviTag plus His_6_ tag. Nanobodies with an N-terminal PelB periplasmic targeting sequence and C-terminal His_8_ tag were generated by inverse PCR and blunt-end ligation of the original clones, generated in pADL-23c. For *in vitro* biotinylation of nanobodies, an AviTag sequence was added ahead of a His_9_ tag using Gibson assembly. For AlexaFluor568 conjugation, a cysteine residue was added before the His_8_ tag by QuikChange mutagenesis. For expression of nanobody-Fc fusions, VHH sequences were cloned into pHLSec [29] encoding the heavy chain of human IgG1, with the VHH domains cloned before the IgG hinge region. For co-translational biotinylation, an AviTag was added to the C terminus of the IgG1 Fc3 domain using inverse PCR and a secreted BirA-FLAG plasmid [30] was used.

### Nanobody generation

Nanobodies were generated as described previously [31]. Briefly, N and Gc spike (200 µg each) were mixed with the adjuvant Gerbu LQ#3000 for each of three intramuscular immunizations of a llama on days 0, 28 and 56. Blood (150 mL) was collected on day 66. Immunizations and handling of the llama were performed under the authority of the project license PA1FB163A. VHHs were amplified by two rounds of PCR from cDNA prepared from peripheral blood monocytes and cloned into the SfiI sites of the phagemid vector pADL-23c (Antibody Design Laboratories, San Diego, USA). Electro-competent *Escherichia coli* TG1 cells (Agilent) were transformed with the recombinant pADL-23c vectors, and the resulting TG1 library stock was infected with M13K07 helper phage to obtain a library of VHH-presenting phage. Phage displaying VHHs specific for N and Gc spike were enriched after two rounds of bio-panning on 50 nM and 5 nM of biotinylated protein respectively, through capturing with Dynabeads M-280 (Thermo Fisher Scientific). After the second round of panning, 93 individual phagemid clones were picked, VHH-displaying phage were recovered by infection with M13K07 helper phage and tested for antigen binding by ELISA with biotin-tagged N or Gc spike immobilised on neutravidin coated plates. Positive phage binders were sequenced and grouped according to CDR3 sequence identity using the IMGT/V-QUEST server [32].

### Mammalian cell culture

Freestyle 293F suspension cells (Thermo Fisher Scientific) were grown in Freestyle 293F medium (Gibco) on a shaking platform (125 rpm) in a humidified 8% CO_2_ atmosphere at 37°C. Cells stably expressing OROV Gc Spike and CRIV Gc Spike under control of the tetracycline response element promoter were generated using the PiggyBac transposase system as previously described [28]. Briefly, A 30 mL suspension culture of Freestyle 293F cells at 1×10^6^ cells/mL was transfected with a 5:1:1 mass ratio of PB-TSW(Gc spike): PB-RN: PBase (35 μg total DNA) using Freestyle MAX transfection reagent (Invitrogen) as per the manufacturer’s instructions. After 2 days, the cells were transferred to fresh media supplemented with 500 μg/mL geneticin (Gibco), and the drug selection was continued for 2 weeks with media replenishment every 3 days.

HeLa and Vero CCL-81 cells were obtained from ATCC (American Type Culture Collection) and cultured in Dulbecco’s Modified Eagle Medium (DMEM; Thermo Fisher Scientific) supplemented with 10% (w/v) foetal bovine serum (FBS), 100 U/mL penicillin and 100 μg/mL streptomycin at 37°C in a humidified 5% CO_2_ atmosphere.

### Protein expression

OROV N was expressed in *E. coli* T7 Express *lysY*/*I^q^*cells (New England Biolabs) in 2×TY medium at 37°C to an OD_600_ of 0.6. The temperature was dropped to 22°C and protein expression by addition of 0.4 mM isopropyl β-D-thiogalactopyranoside (IPTG). The following morning cells were harvested by centrifugation and stored at -80°C until required.

For OROV and CRIV Gc Spike, stably transfected cells at 0.8–1.0×10^6^ cells/mL were induced by addition of 0.5–2 µg/mL doxycycline and cultured for 48–72 h before harvesting of cell pellets. Supernatant was 0.2 µm filtered and stored at 4°C until required.

Nanobodies were expressed in *E. coli* WK6 cells (ATCC) in Terrific Broth supplemented with 2% (v/v) glucose at 37°C to an OD_600_ of 0.8–1.2. Protein expression was induced by adding 1 mM ITPG and the temperature was lowered to 28°C. The following morning cells were harvested by centrifugation and used immediately for protein purification.

For nanobody-Fc fusions, Freestyle 293F cells at 1×10^6^ cells/mL were transiently transfected by addition of DNA that had been pre-incubated in PBS with 25 kDa branched polyethylenimine (PEI), adding 1 μg DNA and 1.5 μg PEI per ml of cultured cells. For nanobody-Fc fusions with AviTags, the plasmid encoding secreted BirA-FLAG was co-transfected with the plasmid encoding the nanobody-Fc fusion at 1:9 DNA ratio and the culture was supplemented with 100 µM D-biotin. Cells were cultured for a further 48 h before cells were harvested. Supernatant was 0.2 µm filtered and stored at 4°C until required.

### Protein purification

For OROV N, pellets were resuspended at 4°C in Ni-NTA lysis buffer (25 mM Tris pH 7.5, 500 mM NaCl, 20 mM imidazole, 0.5 mM MgCl_2_, 1.4 mM β-mercaptoethanol, 0.05% TWEEN-20) supplemented with 200 U bovine DNase I (Merck), 200 μL EDTA-free protease inhibitor cocktail (Merck) and 500 U benzonase (Merck) before lysis using a TS series cell disruptor (Constant Systems) at 24 kpsi. Lysates were cleared by centrifugation (40,000×g, 30 min, 4°C) and incubated with Ni-NTA agarose (Qiagen) for 1 h at 4°C before extensive washing (≥ 20 column volumes [c.v.]) of Ni-NTA wash buffer (20 mM Tris pH 7.5, 500 mM NaCl, 20 mM imidazole) and elution using Ni-NTA elution buffer (20 mM Tris pH 7.5, 500 mM NaCl, 250 mM imidazole).

For OROV and CRIV Gc spike, the filtered supernatant was supplemented with 25 mM Tris pH 8.5, 300 mM NaCl and 0.5 mM MgCl_2_ before being incubated with Ni-NTA agarose (Qiagen) for 1 h at 4°C. The Ni-NTA agarose was washed with ≥ 10 c.v. of 20 mM Tris pH 8.0, 300 mM NaCl and proteins were eluted using PBS supplemented with 250 mM imidazole pH 7.5.

For nanobodies, the pellet from 1 L of bacterial culture was resuspended in 15 mL cold TES buffer (200 mM Tris pH 8.0, 500 mM sucrose, 0.5 mM EDTA) before being diluted 3-fold in cold TS buffer (50 mM Tris pH 8.0, 125 mM sucrose) supplemented with 200 U bovine DNase I (Merck) and 200 μL EDTA-free protease inhibitor cocktail (Merck) to lyse the periplasm. After 30 min incubation at 4°C, cells were harvested by centrifugation at 30,000×g. The supernatant was diluted 5-fold with cold PBS supplemented with 0.5 mM MgCl_2_ before being incubated with Ni-NTA agarose for 30–60 min at 4°C. The Ni-NTA agarose was washed with ≥ 10 c.v. of PBS supplemented with 10 mM imidazole pH 7.5 and nanobodies were eluted using PBS supplemented with 250 mM imidazole pH 7.5.

For nanobody-Fc fusions, the supernatant was stabilised by addition of a 100 mM sodium phosphate pH 7.0 at a 6:1 supernatant:buffer ratio. The supernatant was then injected onto HiTrap Protein G or Mab select Prism A columns (Cytiva) that were washed with 20 mM sodium phosphate pH 7.0 before protein was eluted using 0.1 M glycine pH 2.7. The pH of fractions was immediately neutralised via elution into collection vials containing 1 M Tris pH 8.5.

All proteins were subsequently subjected to size-exclusion chromatography using Superdex 75 16/600 (bacterial expression) or 10/300 (mammalian expression) columns (Cytiva) equilibrated with phosphate-buffered saline (PBS).

### Protein labelling

With the exception of nanobody-Fc fusions, proteins with an AviTag were biotinylated *in vitro* using GST-tagged *E. coli* BirA as described in *[33]*. Briefly, 100 µM AviTagged nanobody was incubated with 5 mM MgCl_2_, 2 mM ATP, 150 µM D-biotin and 1 µM GST-BirA at 30°C for 1 h. The reaction was supplemented with a further 150 µM D-biotin and 1 µM GST-BirA before incubation at 30°C for a further 1 h. The GST-BirA was removed by depletion using glutathione Sepharose 4B (Cytiva) and excess biotin was removed via dialysis. The extent of biotinylation was assayed via a streptavidin shift assay, where 0.5, 1 and 2-fold molar excess of purified streptavidin (Leinco Technologies) was added to samples of biotinylated protein that had been boiled in SDS-PAGE loading buffer. The electrophoretic mobility of streptavidin-bound biotinylated nanobody is dramatically altered in SDS-PAGE. This allowed determination of percentage biotinylation, via calculating the ratio of unshifted protein in samples lacking streptavidin versus samples with 2-fold molar excess of streptavidin.

For AlexaFluor568 labelling, 500 µg of Cys-His_8_ tagged nanobodies in 100 µL PBS were supplemented with 2.5 mM TCEP and incubated at 4°C for 10 min before addition of 8.2 mM AlexaFluor568-Maleimide (ThermoFisher Scientific). The reaction was incubated in the dark at 4°C for 1 h before unbound dye was removed using Zeba dye and biotin removal spin columns (ThermoFisher Scientific) according to the manufacturer’s instructions. AlexaFluor-labelled nanobodies were diluted to 1 mg/mL with PBS supplemented with 0.02% sodium azide and stored at 4°C.

Horseradish peroxidase (HRP) conjugation was performed using the EZ-Link plus activated peroxidase kit (Thermo Fisher Scientific) according to the manufacturer’s instructions. FcSpB6 and FcSpC7 were conjugated in PBS, pH 7.4 using a 2:1 molar ratio of activated HRP to nanobody-Fc fusion. FcNpD2 was conjugated in 0.2 M carbonate-bicarbonate buffer, pH 9.4, using a 4:1 molar ratio of HRP to nanobody-Fc fusion. Following HRP conjugation, nanobody-Fc fusions were exchanged into PBS using a PD minitrap G-10 column (Cytiva), mixed 1:1 with 100% (v/v) glycerol and stored at -20°C.

### Virus preparation

Unless otherwise indicated, the OROV strain used in this study was BeAn19991, kindly provided by Prof. Luiz Tadeu Moraes Figueiredo (University of São Paulo). For animal infection, ELISA and *in vitro* neutralisation assays, OROV BeAn19991 [25,34], TRVL [3] and AM0088 [12] stocks were prepared in Vero CCL-81 cells, which were infected when they reached 90 to 95% confluence. The cultures were maintained until approximately 80% of the cells exhibited cytopathic effect. The supernatant was then collected and clarified by centrifugation at 5000×g for 10 minutes at 4°C. Titration was performed in Vero CCL-81 cells infected with serial 10-fold dilutions of the stocks, which were maintained in semi-solid medium containing 0.75% carboxymethyl cellulose (CMC). After 72 h the cells were fixed with 4% (v/v) formaldehyde and stained with 1% (w/v) crystal violet for plaque counting. The titers obtained were 9.0×10^6^ PFU/mL for the BeAn19991 strain, 1.4×10^5^ pfu/mL for TRVL, and 1.0×10^6^ PFU/mL for the AM0088 reassortant.

For immunocytochemistry and immunoblotting, virus stocks were produced via intercranial injection of neonatal C57BL/6 mice. The brain was homogenised when neurological sequelae became apparent, 2–3 days post-infection. For OROV BeAn19991, the homogenate was used to inoculate Vero CCL-81 cells. At 36 hours post-infection (hpi), Vero cell supernatants were clarified by centrifugation (3000×g for 10 minutes), frozen and stored at -80°C. For OROV AM0088, the homogenate was diluted in DMEM and used directly for infections.

### Animals and infections

Animal experiments were conducted according to the ethical standards of the local ethics committee (CETEA) of the Ribeirão Preto School of Medicine, University of São Paulo, under protocol number 1366/2024R1. For monitoring immune response, six-week-old male C57BL/6 mice were infected subcutaneously with 10^6^ PFU of OROV, and blood was collected by facial vein puncture on days 7, 14, and 21 post-infection. For immunizations, 6-week-old male C57BL/6 mice were immunised with intramuscular injections containing 35 µg of OROV Gc spike or a combination of 35 µg OROV Gc spike and 30 µg OROV N, both with incomplete Freund’s adjuvant. Three immunizations were administered, with two-week intervals between them. Prior to the booster immunization, and 6 weeks after the final boost, a total blood sample was collected by puncture of the facial vein. The serum was separated by centrifugation at 3000 rpm for 10 min at room temperature before being used for neutralization and ELISA assays.

### Enzyme-linked immunosorbent assays (ELISAs)

All ELISA assays were performed using 96-well clear flat bottom polystyrene high bind microplates (Corning 9018) using PBS plus 0.05% TWEEN-20 (PBS-T) as wash buffer and PBS-T supplemented with 2% (w/v) bovine serum albumin (BSA) as blocking and antibody/antigen dilution buffer. Plates were washed at least 4 times with PBS-T between incubation or detection steps. Assays were developed using 150 µg/mL 3,3′,5,5′-tetramethylbenzidine (TMB), 0.05% H_2_O_2_ in 0.1 M sodium acetate pH 5.0 for 30 min before mixing with an equal volume of 0.16 M sulfuric acid and measuring absorbance at 450 nm. All incubations were performed at room temperature unless otherwise stated.

For indirect ELISAs to monitor for antibodies against OROV mice, microplates were coated with purified antigen (2 µg/mL in PBS) overnight at 4°C. A standard curve was generated by coating the plates with increasing concentrations of purified mouse IgG (Merck I5381) or IgM (Thermo Fisher Scientific MA1-10438). Plates were blocked for 2 h before being incubated with serum diluted in blocking buffer (1 in 250,000 for IgG and 1 in 200,000 for IgM [infected animals], or 1 in 800,000 for IgG [immunised animals]). Plates were subsequently incubated with isotype-specific HRP-conjugated secondary antibodies (goat anti-mouse IgG, 1:10,000 dilution, Thermo Fisher Scientific 31430, and goat anti-mouse IgM, 1:2,000 dilution, Merck AP128P) for 30–60 min before developing.

For indirect ELISA to assess binding of biotinylated nanobodies to purified antigens, microplates were coated with purified antigen (OROV N, OROV Gc spike or CRIV Gc spike; 10 µg/mL in PBS) for 2 h and blocked overnight. Plates were incubated with biotinylated nanobodies (2 µg/mL in blocking buffer) for 1 h, HRP-conjugated streptavidin (1:10,000 dilution, Thermo Fisher Scientific N100) for 15 min, and then developed.

For sandwich ELISAs using nanobodies, microplates were coated with His_8_-tagged nanobodies (2 µg/mL in PBS) for 2 h and blocked overnight at room temperature or 4°C. Plates were then incubated with purified antigens in blocking buffer (OROV N or OROV Gc spike, 10 µg/mL or diluted as shown for standard curves, or virus stocks diluted as shown) for 1 h, biotinylated nanobody (2 µg/mL in blocking buffer) for 30–60 min, HRP-conjugated streptavidin (1:10,000 dilution) for 15 min, and then developed.

For sandwich ELISAs using nanobody-Fc fusions, microplates were coated with neutravidin (10 µg/mL in PBS, Fisher Scientific A2666) for at least overnight at 4°C. Plates were incubated with capture reagent (biotinylated nanobody-Fc fusion, 2 µg/mL in blocking buffer) for 1 h, blocked for 1 h, incubated with antigen in blocking buffer (serum diluted 1 in 4, virus stocks diluted as shown, or purified antigens diluted as shown for standard curves) overnight at 4°C, incubated with detection reagent (HRP-conjugated nanobody-Fc fusion, 0.4 µg/mL in blocking buffer), then developed.

For calculating limits of detection for sandwich ELISAs, standard curves of antigen were generated using purified antigen and the standard deviation of the background was calculated from at least four wells to which no antigen was added. The limit of detection is calculated as 3.3 * standard deviation of background / slope of standard curve [35].

RT-qPCR analysis of clinical samples was performed as described previously [20]. Least squares non-linear regression of the log_10_-transformed Z scores and Ct values was performed using Prism 7 (GraphPad Software).

### Neutralization assays

For neutralization using nanobody-Fc fusions or animal serum, the samples were two-fold serially diluted with PBS (from 2 µM for nanobody-Fc fusions and from 1:25 for serum) before being incubated with an equal volume of virus particles for 1 h at 37°C. The inoculum was added to Vero CCL-81 cells at 95% confluence, and the cultures were incubated at 37°C with gentle manual agitation every 15 min. After 1 h, the inoculum was removed and semi-solid medium (DMEM with 2% (w/v) FBS, 100 U/mL penicillin, 100 μg/mL streptomycin, 0.25 μg/mL Amphotericin B and 0.75% CMC) was added. Staining was performed 72 hpi using 1% (w/v) crystal violet, following fixation with 4% (v/v) formaldehyde for 30 min. Plaques were counted and neutralization was assessed by evaluating the infection of OROV at each nanobody concentration relative to a PBS control. The neutralizing dose (*ND_50_*) was calculated by non-linear fitting to a three-parameter inhibitor-response curve where the top and bottom values were constrained to 100 and 0, respectively, using Prism 7 (GraphPad Software).

### Immunocytochemistry

Confluent monolayers of HeLa were infected by addition of OROV diluted in serum free media (MOI 0.5) and rocking the cells on ice for 1 h. Cells were moved to 37°C for 15 min, washed with PBS, and then overlain with DMEM with 2% (w/v) FBS. At 20 hpi cells were washed with PBS and fixed for 10 min at with 4% (v/v) paraformaldehyde in PBS. Cells were permeabilised for 30 min using 0.1% saponin in blocking buffer (0.2% porcine gelatine in PBS). Permeabilised cells were incubated with primary antibodies and anti-OROV nanobodies at 37°C for 30 min, diluted as follows: mouse anti-OROV antiserum, 1:500 [36]; AlexaFluor568 tagged anti-OROV nanobodies, 1:4000; anti-TGN46 (Bio-Rad AHP500), 1:500. After washing, secondary antibodies were applied at 37°C for 30 min, diluted as follows: AlexaFluor488 anti-mouse and AlexaFluor647 anti-sheep (Thermo Fisher Scientific), 1:1000. Cells were mounted on glass slides using Fluoromount-G and imaged at 40× magnification using a Zeiss LSM 780 confocal microscope at the Ribeirão Preto Medical School Multiuser Laboratory of Multiphoton Microscopy.

### Immunoblotting

Confluent monolayers of HeLa cells in 6-well plates were washed with PBS and then infected by addition of OROV (MOI 5) with gentle rocking on ice for 90 min before transfer to 37°C for 15 min to facilitate virus entry. Cells were washed and incubated in DMEM with 2% (w/v) FBS at 37°C. At 24 hpi cells were washed, detached by incubation with 2 mM EDTA in PBS for 5 min, harvested by centrifugation at 2000×g for 10 min at 4°C, and the cell pellets were stored at -80°C. Cells were resuspended and incubated on ice for 20 min in lysis buffer comprising 50 mM Tris pH 7.5, 150 mM NaCl, 10% (v/v) glycerol, 5 mM EDTA, 1% (v/v) Triton X-100, plus protease inhibitors (P8340; Sigma-Aldrich). Lysates were clarified (16,000×g, 10 min, 4°C), protein concentration was equalised following analysis using the Bradford assay (Bio-Rad), and samples were diluted in Laemmli buffer supplemented with β-mercaptoethanol. Samples were heated to 95°C for 5 min, separated by SDS-PAGE and transferred to 0.45 µm nitrocellulose membranes (Millipore). Membranes were blocked using 5% (w/v) skim milk powder in PBS with 0.1% TWEEN-20 for 1 h before being incubated with either mouse anti-OROV antiserum (1:500) then HRP-conjugated sheep anti-mouse (1:1000, Cytiva), or with HRP-conjugated nanobody Fc-fusions FcNpD2, FcSpB6 or FcSpC7 (1:1000). Protein bands were visualised using enhanced chemiluminescence reagents and the ChemiDoc Imaging System with ImageLab software (Bio-Rad).

## Results

Orthobunyaviruses form enveloped particles that enclose an RNA genome tightly bound by the nucleoprotein (N) and possess the glycoproteins Gn and Gc on their surface [37]. The N-terminal region of Gc forms a prominent ‘spike’ on the surface of orthobunyavirus particles, composed of conserved ‘head’ and ‘stalk’ domains [38–40]. Previous studies have shown that OROV infection elicits antibodies directed against the proteins N and Gc [41,42], suggesting that both these antigens are accessible to the humoral immune system. OROV N was thus purified following recombinant expression in *E. coli*, and the Gc spike domain (residues 482– 894) was purified following expression using mammalian (Freestyle 293F) cells to allow protein glycosylation (Fig. 1A). Purified OROV N had a 260:280 nm absorbance ratio ≥ 1.5, consistent with purification of N bound to bacterial RNA [43]. Purified antigens were used as capture antigens in an indirect ELISA and incubated with serum from mice experimentally infected with OROV. Both N and Gc spike are recognised by IgG (Fig. 1B) and IgM (Fig. S1A) from serum of infected mice. Neither antigen is recognised by antibodies present in a mock-infected mouse, nor is there recognition of the Gc spike from the related Cristoli orthobunyavirus (CRIV; Fig. 1B and S1A). This confirms that the purified recombinant antigens display authentic epitopes that are presented to the immune system during OROV infection. To test their ability to raise a neutralising immune response, purified OROV antigens (Gc spike with or without N) were used to immunize 6-week-old mice using a prime-boost-boost regimen. Both antigens raised a strong, specific IgG response (Fig. S1B) that was potently able to neutralise OROV infection *in vitro* (Fig. 1D).

**Figure 1.**
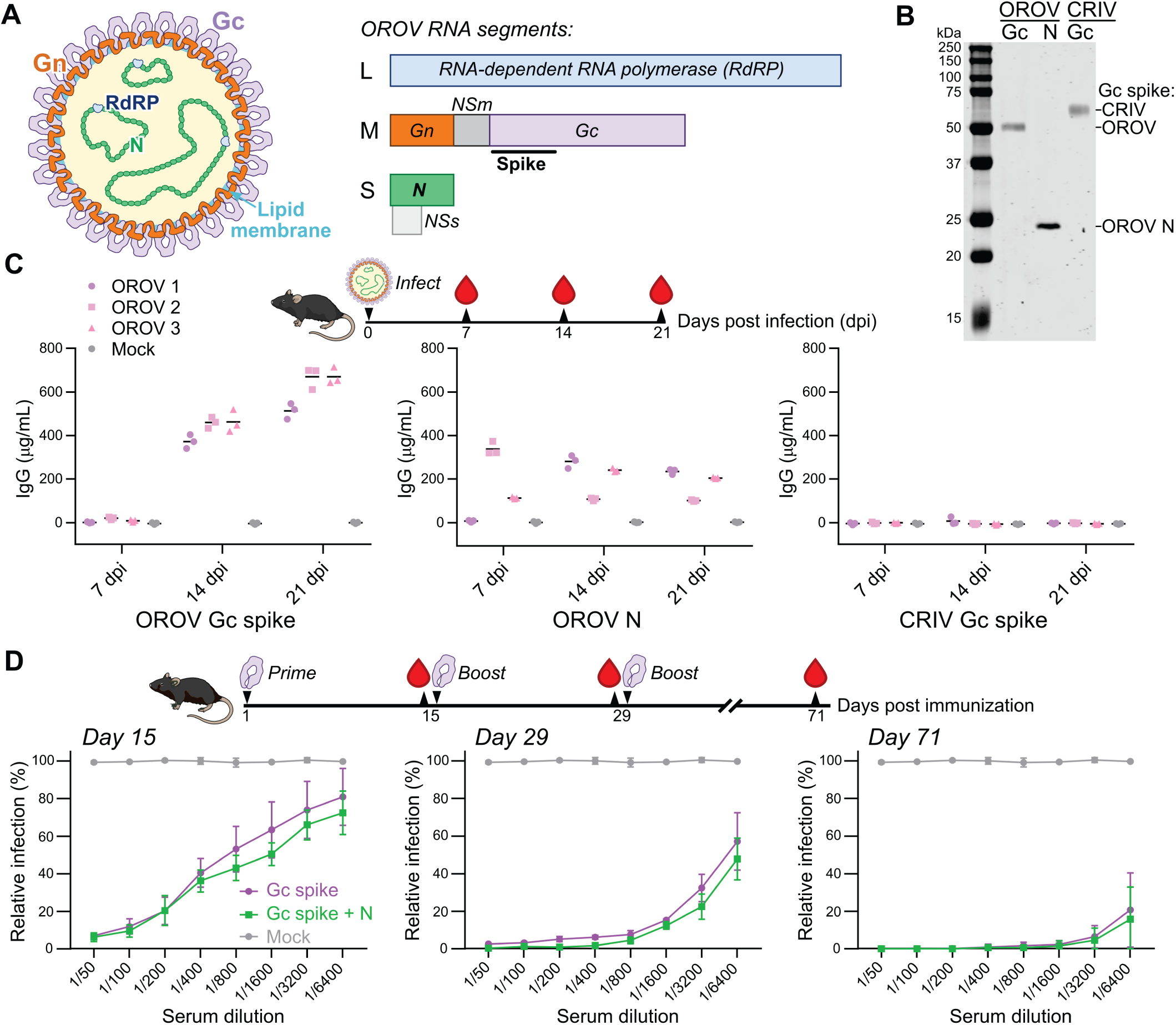
Purification and characterisation of recombinant OROV antigens. (**A**) Schematic representation of OROV virion (*left*) and RNA genome segments (*right*). The RNA-dependent RNA polymerase (RdRP) associates with the circular genome segments and is encoded by the large (L) segment. The surface glycoproteins Gn and Gc, plus the non-structural protein NSm, are encoded by the medium (M) segment polyprotein. The genome-associated nucleoprotein (N) and non-structural protein NSs are encoded by the small (S) segment. Nanobodies were raised against the N and the spike region of Gc (residues 482–894 of the M polyprotein). (**B**) Coomassie-stained SDS-PAGE of purified OROV antigens, and the Gc spike region of the related orthobunyavirus Cristoli virus (CRIV). (**C**) Purified antigens are recognised by antibodies raised by mice in response to infection with OROV strain BeAn19991. Blood was harvested at indicated days post-infection (dpi) and an indirect ELISA to detect IgG was performed using indicated antigens. Values are show for three independent ELISA experiments using serum from three infected mice (OROV 1–3) and one mock-infected control mouse. (**D**) Neutralising antibodies are elicited by immunization with purified OROV antigens. Mice were immunised with OROV Gc spike alone, or Gc spike plus N, and blood samples were taken at indicated times. Serum was diluted for plaque reduction neutralization assays, with relative number of plaques compared to the mock-immunised mouse shown. Each point represents data for three mice (mean ± SD).

Purified OROV N and Gc spike were used to immunize a llama and nanobodies were isolated, enriched and cloned following the procedure outlined in [31]. Sequence analysis following enrichment with phage display identified multiple distinct clusters for each antigen, and for each antigen six nanobody sequences that were representative of each unique cluster (Fig. S2A, Table S1) were selected for further characterisation. Nanobodies were cloned and purified following *E. coli* expression with a His_8_ tag alone or with a His_9_ tag plus biotin acceptor peptide (AviTag) to facilitate *in vitro* biotinylation (Fig. S2B–D), and indirect ELISA experiments confirmed that the biotinylated nanobodies could recognise purified antigen (Fig. 2A). Sandwich ELISA experiments with purified antigens, performed using passive adsorption of the His_8_-tagged capture nanobody and detection of the biotinylated nanobody via horseradish peroxidase (HRP)-conjugated streptavidin, identified two competition groups for the Gc spike nanobodies, with one group composed of nanobodies SpB6, SpB7 and SpE8 while the second comprised only SpC7 (Fig. 2B). We did not observe any competition for epitopes in the N protein sandwich ELISA (Fig. 2B). Further characterisation identified SpB6:(biotinylated)bSpC7 and NpF2:bNpE3 as the optimal pairings for capture: detection of Gc spike and N, respectively, able to detect picogram quantities of purified OROV antigens (Fig. 2C). These nanobodies recognise laboratory-grown stocks of OROV strains BeAn19991 and TRVL, with the N nanobodies demonstrating higher sensitivity (Fig. 2D). However, preliminary experiments demonstrated that sandwich ELISAs using these nanobodies were insufficiently sensitive to detect OROV antigens in serum samples from human patients with acute OROV infection.

**Figure 2.**
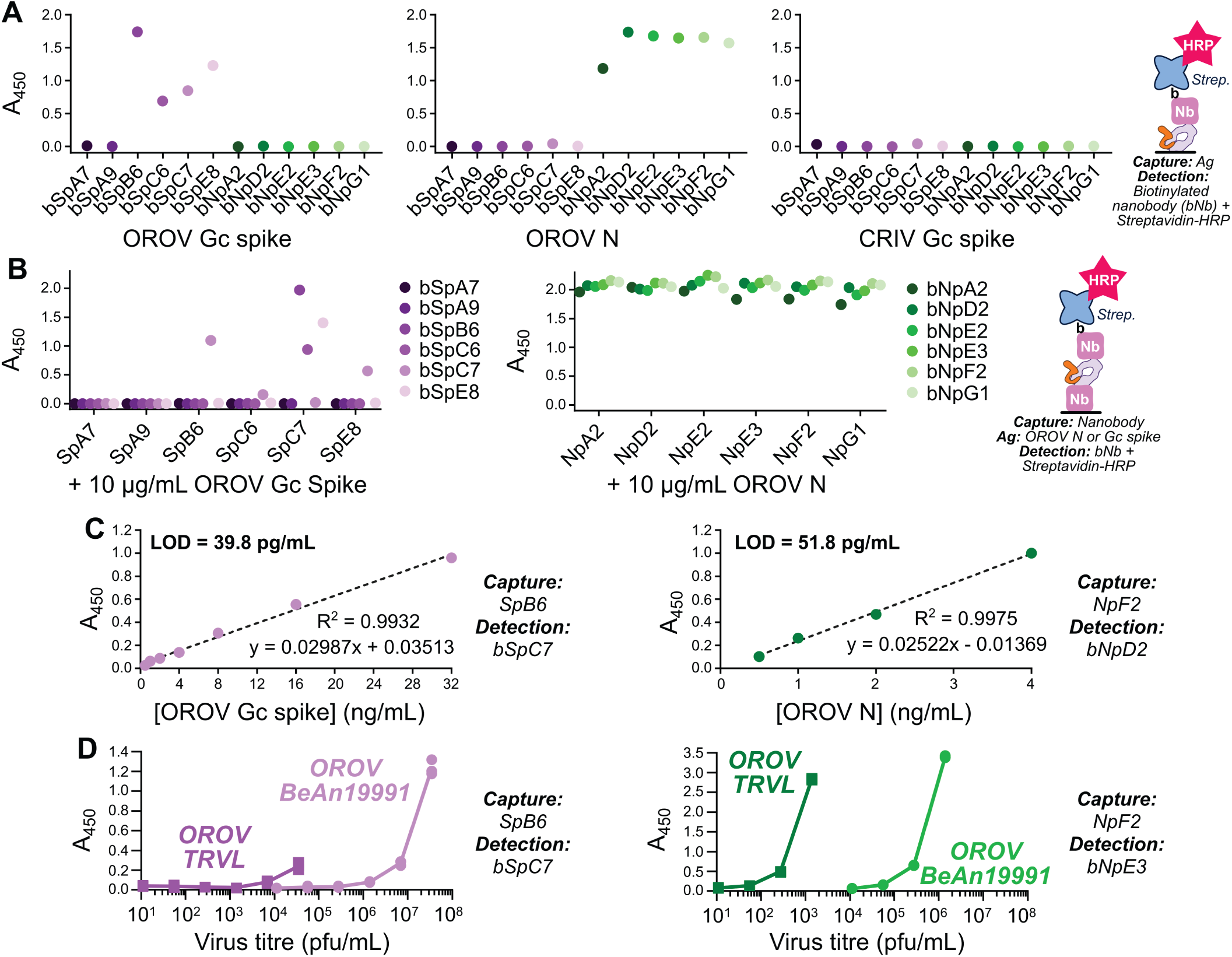
Nanobodies for specific detection of OROV Gc spike and. **N.** (**A**) Nanobody detection of purified OROV antigens. Antigens were immobilised on the capture surface via passive adsorption (2 µg/mL) and detected via incubation with specified biotinylated nanobodies (bNbs) plus streptavidin-HRP. CRIV Gc spike was included as a negative control. (**B**) Sandwich ELISA using purified OROV antigens to determine nanobody competition groups. Surfaces were coated with nanobodies (2 µg/mL) before being incubated with antigen then detection bNbs and streptavidin-HRP. Data is representative of at least 2 independent experiments. (**C**) Limit of detection (LOD) of purified Gc spike (*left*) or N (*right*) in sandwich ELISA using optimised nanobody pairings. LOD is calculated as (3.3 * standard deviation [background]) / slope. (**D**) Detection of Gc spike (*left*) and N (*right*) in laboratory stocks of OROV strains TRVL [3] and BeAn19991 [25,34] by sandwich ELISA.

Multiple parallel improvements were attempted to enhance sandwich ELISA sensitivity. The nanobodies were cloned as fusions with the linker region plus Fc domains 2 and 3 of human IgG1, thereby generating bivalent detection reagents resembling heavy-chain only antibodies. The resultant dimerization, driven by the IgG1 domains, was anticipated to confer avidity-enhanced antigen binding [35,44]. In addition to untagged constructs, these nanobody-Fc fusions were cloned with a C-terminal AviTag to facilitate co-translational biotinylation, allowing directional capture on ELISA plates coated with neutravidin that should enhance exposure of the antigen-binding motifs. Additionally, the detection nanobody-Fc fusions were covalently coupled to the HRP enzyme. For the sake of expediency only one nanobody-Fc fusion was generated for the N protein (FcNpD2), while two nanobody-Fc fusions were generated Gc spike directed nanobodies (FcSpC7 and FcSpB6; Fig. S3). Of these, the bFcNpD2:FcNpD2-HRP combination proved the most sensitive in detecting purified OROV antigens in a sandwich ELISA (Fig. 3A) with a limit of detection (LOD) of 11.3 ± 7.4 pg/mL (mean ± SD, n = 5 independent experiments), while the bFcSpC7:FcSpB6-HRP combination had a LOD of 572.1 ± 166.5 pg/mL (n = 3 independent experiments). Sandwich ELISAs using the N nanobody-Fc fusions detect laboratory-grown stocks of OROV (Fig. 3B). Importantly, it also recognises the 2023–4 epidemic isolate (AM0088) [12] of OROV (Fig. 3B). OROV AM0088 N shares 100% amino acid identity with the purified N used for nanobody generation.

**Figure 3.**
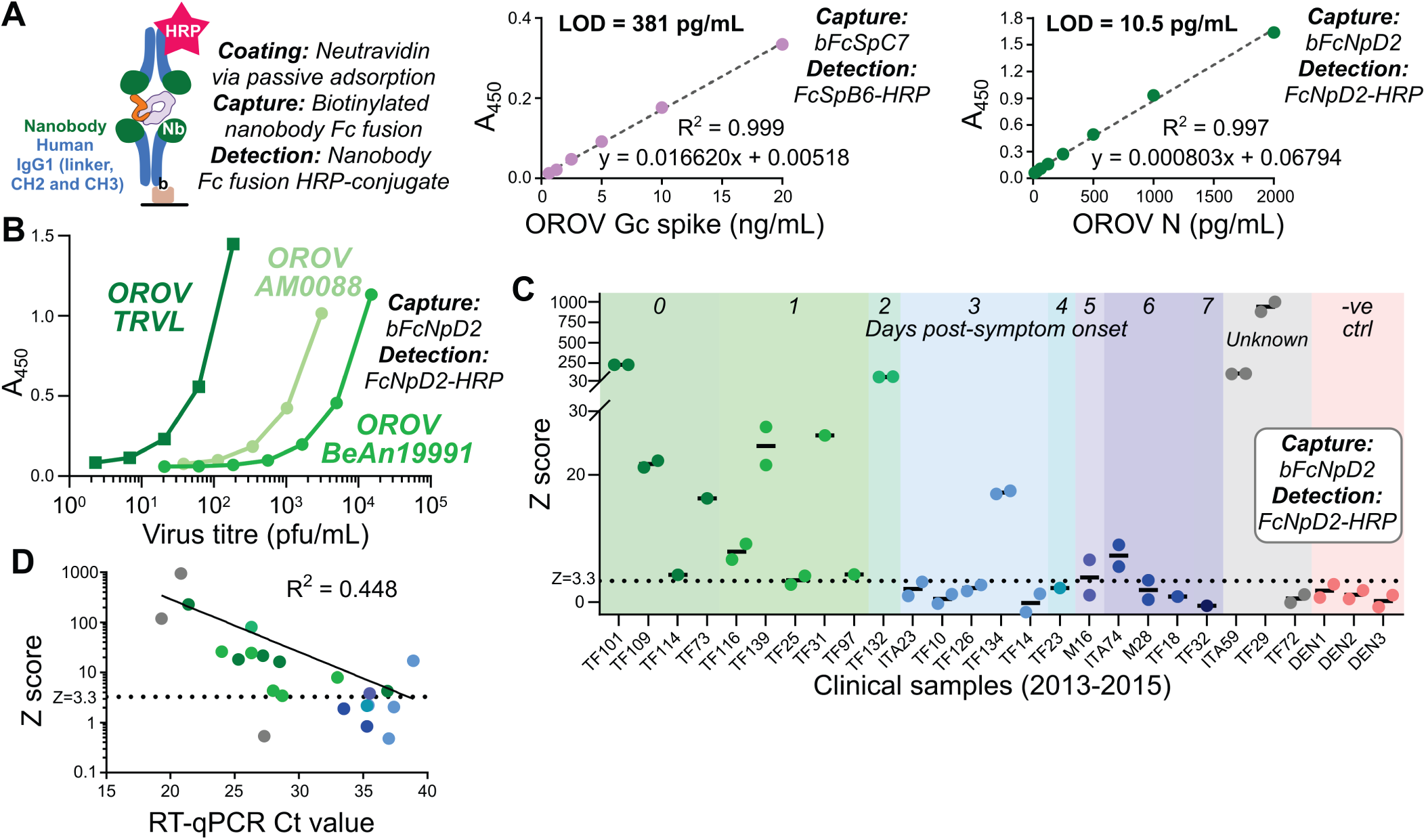
An improved sandwich ELISA allows detection of OROV N in in clinical samples. (**A**) Schematic representation of modified sandwich ELISA (*left*) plus standard curves to determine LOD for purified OROV Gc spike (*middle*) and N (*right*). (**B**) Detection of N in laboratory stocks of OROV strains TRVL [3], BeAn19991 [25,34] and the newly emerged reassortant AM0088 [12] by sandwich ELISA. (**C**) Detection of N in a 1:3 dilution of serum from patients infected with OROV between 2013 and 2015 (Table 1). Antigen values are shown as standard deviations above the mean for blank samples (Z score). PCR-confirmed OROV positive samples are grouped by day of collection post symptom-onset and three DENV-positive samples are shown as negative controls. Data represent 1 or 2 independent measurements for each sample. (**D**) Correlation of Z scores from OROV N sandwich ELISA versus RT-qPCR evaluation of viral genome abundance (Ct values). Non-linear regression analysis of the Ct values and log_10_-tranformed Z scores is shown.

**Table 1.**
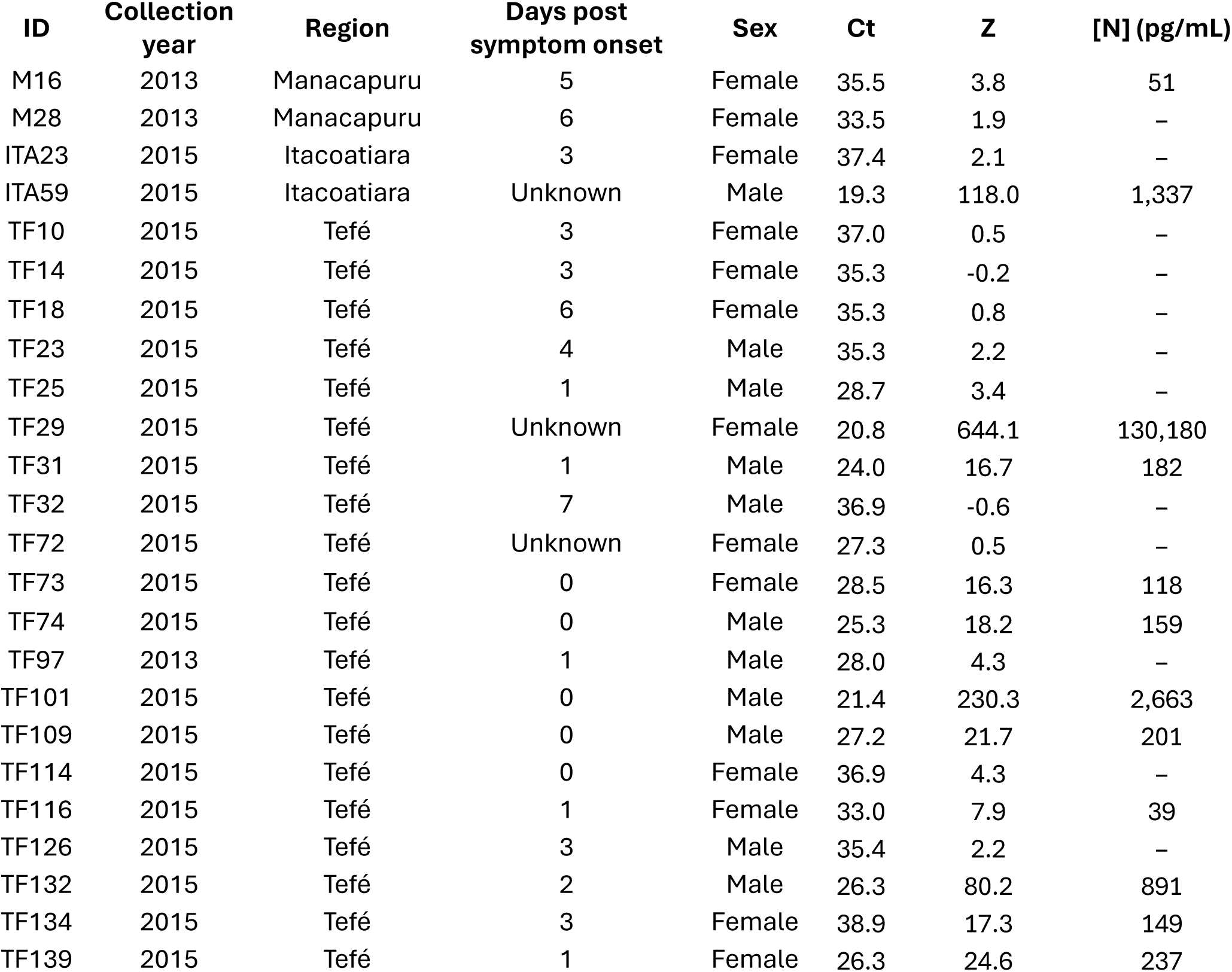
Clinical samples (2013–2015) used in this study. Patients were aged 8 to 65. All were confirmed as positive for OROV by RT-qPCR. Where available, collection time (days post symptom onset) is show. Ct values from RT-qPCR are shown. Average Z score from sandwich ELISA is shown, as is average concentration of N protein where samples were positive (Z ≥ 3.3) and within the range of the antigen standard curve.

The optimised bFcNpD2:FcNpD2-HRP capture: detection sandwich ELISA was used to detect OROV antigens in serum samples that were collected from patients with acute febrile illness between 2013 and 2015 and confirmed as positive for OROV by RT-qPCR (Table 1). OROV N protein was successfully detected above the noise level (Z score ≥ 3.3) for all replicates experiments in 13 of 24 samples (Fig. 3C), with a further 3 samples being alternatively above and below the detection limit in duplicate experiments. No signal above the noise level was detected for three serum samples collected from patients with acute febrile illness that were PCR positive for dengue virus (Fig. 3C). The level of signal in the sandwich ELISA was correlated to the Ct values obtained by RT-qPCR (R^2^ = 0.448; Fig. 3D), with robust detection in the sandwich ELISA for samples with Ct values lower than 26.

In addition to their utility for antigen-based diagnostic assays, nanobody-based reagents are highly suitable for use as research reagents in several formats owing to their ease of expression and purification. Immunoblotting of lysates from OROV-infected cells demonstrate that N is recognised by its cognate HRP-conjugated nanobody-Fc fusion while Gc spike is not (Fig. 4A), suggesting that NpD2 recognises a linear epitope and while SpB6 and SpC7 recognise conformational epitopes. The N and Gc spike nanobodies were purified from *E. coli* following expression with a C-terminal Cys-His_8_ tag, allowing covalent conjugation of fluorescent dyes via maleimide chemistry. Fluorescence microscopy showed that both the N and Gc spike nanobodies can detect OROV antigens in experimentally infected HeLa cells (Fig. 4B). For N the staining substantially overlaps with the signal observed using polyclonal mouse serum obtained from experimentally infected animals, with signal localising primarily to cytoplasmic puncta that may represent virus genome assembly compartments and/or sites of N protein aggregation. There was also some nanobody NpG1 signal in the nucleus of infected cells. For Gc spike there is very limited overlap between nanobody and polyclonal mouse serum signal; nanobody SpE8 signal co-localises with the trans-Golgi marker TGN46, a known site of virus assembly [45], and is observed in small puncta throughout the cell that may represent newly assembled intracellular virus particles. The overlap in signal between nanobody SpE8 and TGN46 is less pronounced in cells infected with OROV AM0088, with signal from SpE8 and polyclonal mouse serum co-localising at large puncta in the periphery of the cells, and there is more pronounced nuclear signal for NpG1 in AM0088-infected cells (Fig. S4).

**Figure 4.**
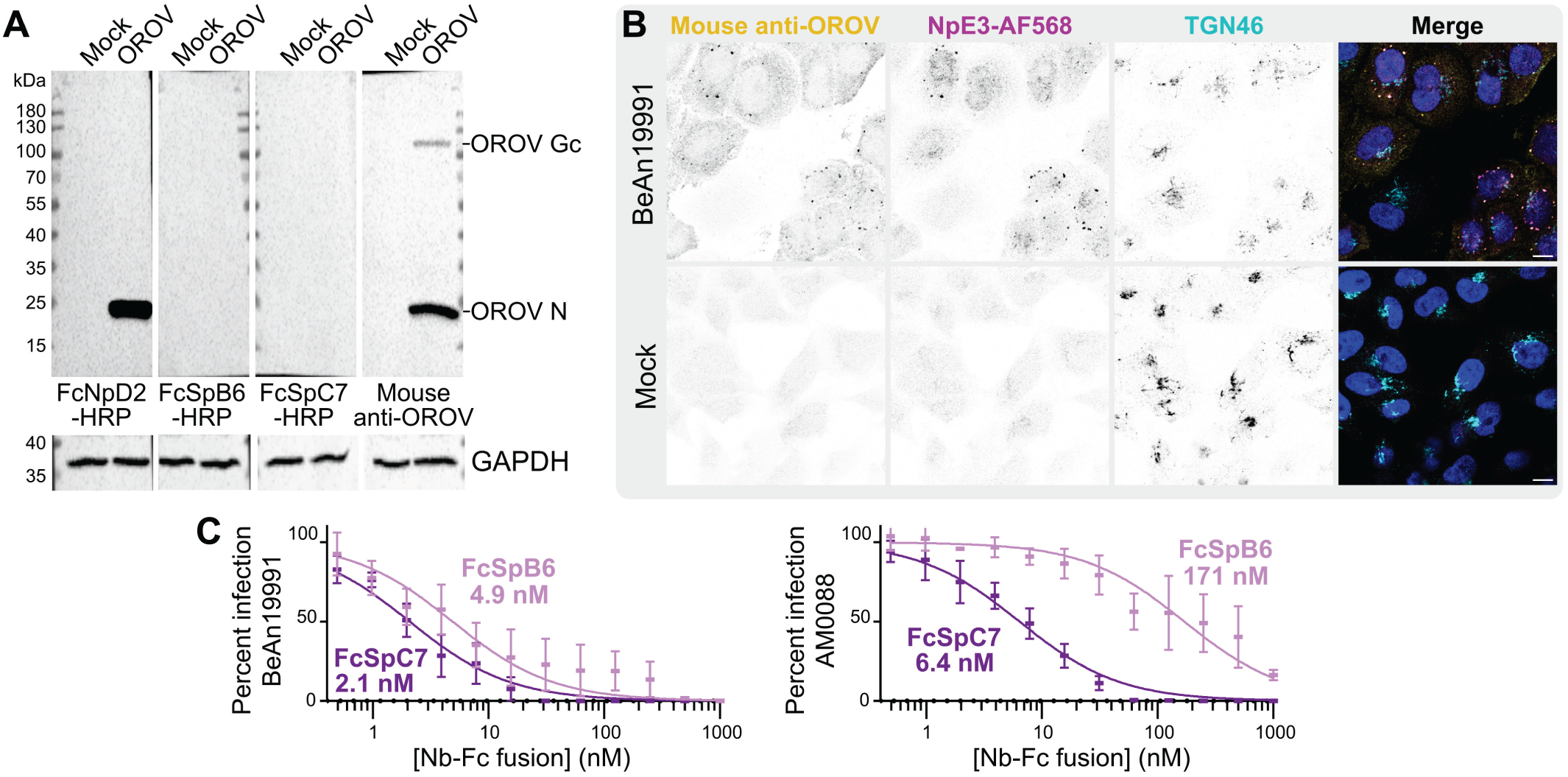
New tools for OROV research. (**A**) Immunoblots of lysates from HeLa cells mock-infected or infected with OROV BeAn19991 (MOI 5). Membranes were probed with HRP-conjugated nanobody-Fc fusions or with a polyclonal antibody against OROV plus anti-mouse secondary antibody, as indicated. Membranes were subsequently probed for GAPDH as a loading control. (**B**) Immunocytochemistry of HeLa cells infected with OROV BeAn19991 (MOI 0.5). Cells were probed with AlexaFluor (AF)568 conjugated OROV nanobody NpE3, with a polyclonal antibody against OROV and with an antibody against TGN46, plus appropriate secondary antibodies. Nuclei are stained with DAPI (blue). Scale bar = 10 µm. (**C**) Nanobody-Fc fusions that recognise Gc spike neutralise OROV infection *in vitro*. Plaque reduction neutralisation assay using Gc spike Nb-Fc fusions, performed using OROV BeAn19991 (top) or AM0088 (bottom). Mean ± SD and *ND_50_*values are shown (n=3 independent experiments).

Infection of HeLa cells with OROV BeAn19991 following incubation with nanobody-Fc fusions demonstrates that FcSpB6 and FcSpC7 neutralise infection with 50% neutralising dose (*ND_50_*) values of 4.9 nM ± 0.79 and 2.1 ± 0.18 nM (*ND_50_* ± standard error, n=3 independent titrations), respectively (Fig. 4C). FcSpB6 has substantially reduced ability to neutralise infection by the new epidemic AM0088 isolate (*ND_50_* = 171 ± 26.3 nM), whereas FcSpC7 retains potent neutralisation ability (*ND_50_* = 6.4 ± 0.54 nM).

## Discussion

Here we present a sandwich ELISA that allows detection of the N protein in the serum of patients suffering from acute OROV infection. While multiple groups have published IgM capture or indirect ELISAs to detect OROV antibodies [41,46–48], to the best of our knowledge this is the first demonstration of a sandwich ELISA to detect OROV antigens in clinical samples.

The efficiency of detection was much higher in samples collected within the first three days following symptom onset (Fig. 3C). While the signal in the sandwich ELISA is correlated with viral genome abundance as measured by RT-qPCR (Fig. 3D), this assay is currently less sensitive. Sensitivity of the ELISA assay could be improved by using two distinct anti-N protein nanobody-Fc fusions for capture and detection, or by engineering these reagents to encode multiple nanobodies fused to the Fc region to enhance the avidity of binding. Having established proof-of-principle that antigen detection can be used for OROV molecular diagnosis, it should be possible to develop a nanobody-based lateral flow device for diagnosing OROV infection at primary points-of-care. Rapid point-of-care diagnosis of OROV would enhance surveillance efforts, which could be especially important for pregnant women in endemic areas, and it would dramatically accelerate enrolment of patients into trials aimed at determining post-infection sequelae or the efficacy of future therapies.

OROV is a member of the Simbu serogroup, the N proteins of which share high amino acid identity (>60%) with OROV N [49]. Serological tests suggest that there is significant cross-reactivity in the antibody response to N proteins, with OROV N being detected by antisera raised against multiple different viruses in the Simbu serogroup [41]. We have not investigated the cross-reactivity of our nanobodies with other Simbu serogroup N proteins. In a clinical setting, it would be prudent to use an orthogonal molecular technique like RT-PCR to confirm and further characterise samples where OROV antigens are detected by the ELISA presented here.

The purified antigen used for immunisation of llamas to produce nanobodies was also tested for its ability to elicit neutralising antibody responses in mice. Immunisation with either OROV Gc spike alone, or Gc spike plus N, raised a strong neutralising antibody response with a 1/6400 dilution of serum taken 6 weeks after the final boost retaining the ability to reduce infection *in vitro* by ∼80% (Fig. 1D). This compares favourably to the only other published immunisation trial for OROV, which used a vesicular stomatitis virus pseudotype system expressing the full OROV M segment or a variant with the Gc head domain removed [50]. Immunisation with the VSV pseudotypes protected mice from clinical symptoms of OROV infection but not from viraemia [50]. Previous studies of Schmallenberg virus (SBV), a related ruminant orthobunyavirus, have shown that immunisation using the Gc head domain or Gc spike (head+stalk) is sufficient to protect mice against lethal challenge [39,51]. Furthermore, the majority of neutralising antibodies raised by SBV infection recognise the head domain of Gc [39]. Recombinant OROV Gc spike is thus a promising candidate for further vaccine studies using protein or mRNA-based immunisation.

Fc fusions of nanobodies that recognise Gc spike are capable of neutralising OROV infection (Fig. 4C), with FcSpC6 potently neutralising both BeAn19991 and the newly emerged AM0088 reassortant (*ND_50_* = 2.1 and 6.4 nM, respectively). Neutralising monoclonal antibodies that recognise the SBV Gc head less potently (*ND_50_* = 0.15–1.8 µM) are sufficient to protect mice from lethal challenge [39]. This suggests that our Gc spike nanobody-Fc fusions could be used as direct-acting antiviral compounds to prevent or treat OROV infection, although confirmation of this requires further study.

The nanobodies raised against OROV Gc spike comprise two competition groups (Fig. 2B), consistent with previous reports that the spike region of OROV Gc is monomeric in solution [39]. The cell surface receptor Lrp1 has recently been identified mediating OROV infection, likely via binding to the OROV Gn protein [52]. The surface glycoproteins of orthobunyavirus particles undergo dramatic structural rearrangement during viral endocytosis, which may prime the particles for cell entry [40]. It will be interesting in the future to define the molecular basis of neutralisation by our Gc spike nanobodies, to determine whether they prevent infection by inhibiting a required conformational change, via steric hindrance of Lrp1 binding, or by preventing binding of an additional as-yet unidentified co-receptor.

Our recombinant N protein was contaminated with nucleic acid, as evidenced by the high 260:280 nm absorbance ratio. This is consistent with previous reports that OROV N co-purifies with bacterial RNA when expressed in *E. coli* [43]. OROV N forms polymers when encapsidating viral genomes *in vivo*. Given that OROV N will be bound to RNA in virus particles and infected cells, we considered RNA-bound N to be representative of the antigen likely to be found in patient serum and encountered by the humoral immune system. We also observed that nanobodies directed at N do not compete for binding (Fig. 2B). Given the small size of OROV N (26.3 kDa) it is unlikely that all six nanobodies bind distinct sites on the protein. Instead, the lack of competition probably arises from the propensity of OROV N purified from *E. coli* to form oligomers [43].

In immunocytochemistry, there was extensive co-localisation between the anti-N nanobody NpE3 and the signal from an immune serum raised in mice (Fig. 4B). However, compared with immune serum we observed more extensive nuclear staining with NpE3. This could be a reflection of different epitopes recognised by the immune serum versus NpE3, or the ability of nanobodies to better access epitopes in crowded environments owing to their small size [53]. A recent study characterised two novel monoclonal antibodies (mAbs) raised against OROV N by immunisation of mice with OROV infected-cell supernatant [23]. They demonstrated the utility of their OROV mAbs for immunocytochemistry following experimental infection of several different cell types, and for immunohistochemistry in experimentally infected animals, but they did not report characterisation of the N protein subcellular localisation. The differences in NpE3 staining observed between BeAn19991 and AM0088 (Figs 4B, S4), despite the N protein being 100% identical across these two species, suggests differences in the virus lifecycle that should be further investigated.

In summary, we have developed protein-based reagents for the detection and characterisation of OROV infection. Purified OROV antigens enable indirect ELISA serology and show promise as candidate vaccine molecules. Nanobodies that recognise OROV Gc spike and N are valuable research reagents. Fc fusions of nanobodies against N enable detection of OROV antigens in serum samples, opening the way to development of low-cost antigen detection tests, and nanobody-Fc fusions that recognise Gc spike potently neutralise infection by both historical and recently emerged isolates of OROV. The reagents presented here will accelerate laboratory and clinical research into OROV, an important emerging pathogen.

## Acknowledgements

We thank Bernard Kelly (University of Cambridge) for the pMWCAVI plasmid and DNA encoding human IgG1, and Prof. Luis Tadeu Moraes Figueiredo (University of São Paulo) for the kind gifts of OROV BeAn19991, TRVL and anti-OROV mouse antiserum. This work was supported by a Biotechnology and Biological Sciences Research Council (BBSRC) grant to RJO (BB_V018523), an International Collaboration Award from the Royal Society to EA and SCG (ICA\R1\201019), a São Paulo Research Foundation (FAPESP) grant to EA (19/26119-0), and International Science Partnerships Fund (ISPF) Institutional Support Grant (ODA) from Research England to SCG. The funders had no role in study design, data collection and analysis, decision to publish, or preparation of the manuscript. For the purpose of open access, the author has applied a Creative Commons Attribution (CC BY) licence to any Author Accepted Manuscript version arising.

## Ethics statement

Immunizations and handling of llama were performed under the authority of UK Home Office project license PA1FB163A. All mouse experiments were approved by the local ethics committee at the Ribeirão Preto School of Medicine (CETEA), under protocol number 1366/2024R1. Use of human clinical samples was approved by the Fundação de Medicina Tropical Dr. Heitor Vieira Dourado ethics committee under registration number 700.915.

## Data availability

The authors confirm that the data supporting the findings of this study are available within the article and its supplementary materials. Plasmids encoding the proteins described in this study, and selected cell lines plus purified proteins, will be made available from the Centre for Infectious Disease Reagents (https://nibsc.org/science_and_research/idd/cfar.aspx), UK Medicines and Healthcare products Regulatory Agency, upon publication.

---

**Table S1.**
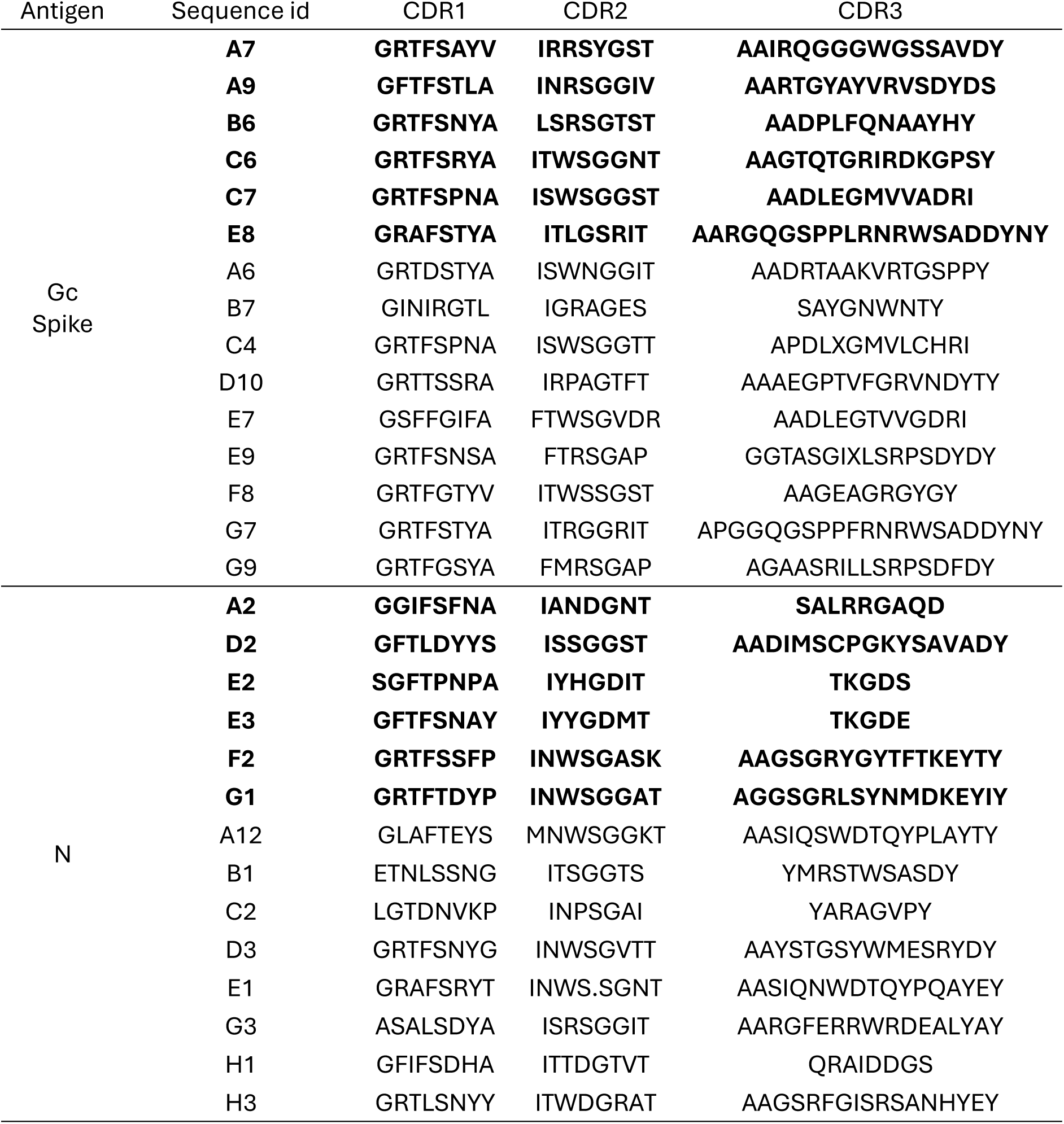
Nanobody sequences. The complementarity determining region (CDR) of the unique nanobody (VHH) sequences isolated by phage display are shown, with selected nanobodies highlighted in **bold**.

**Figure S1.**
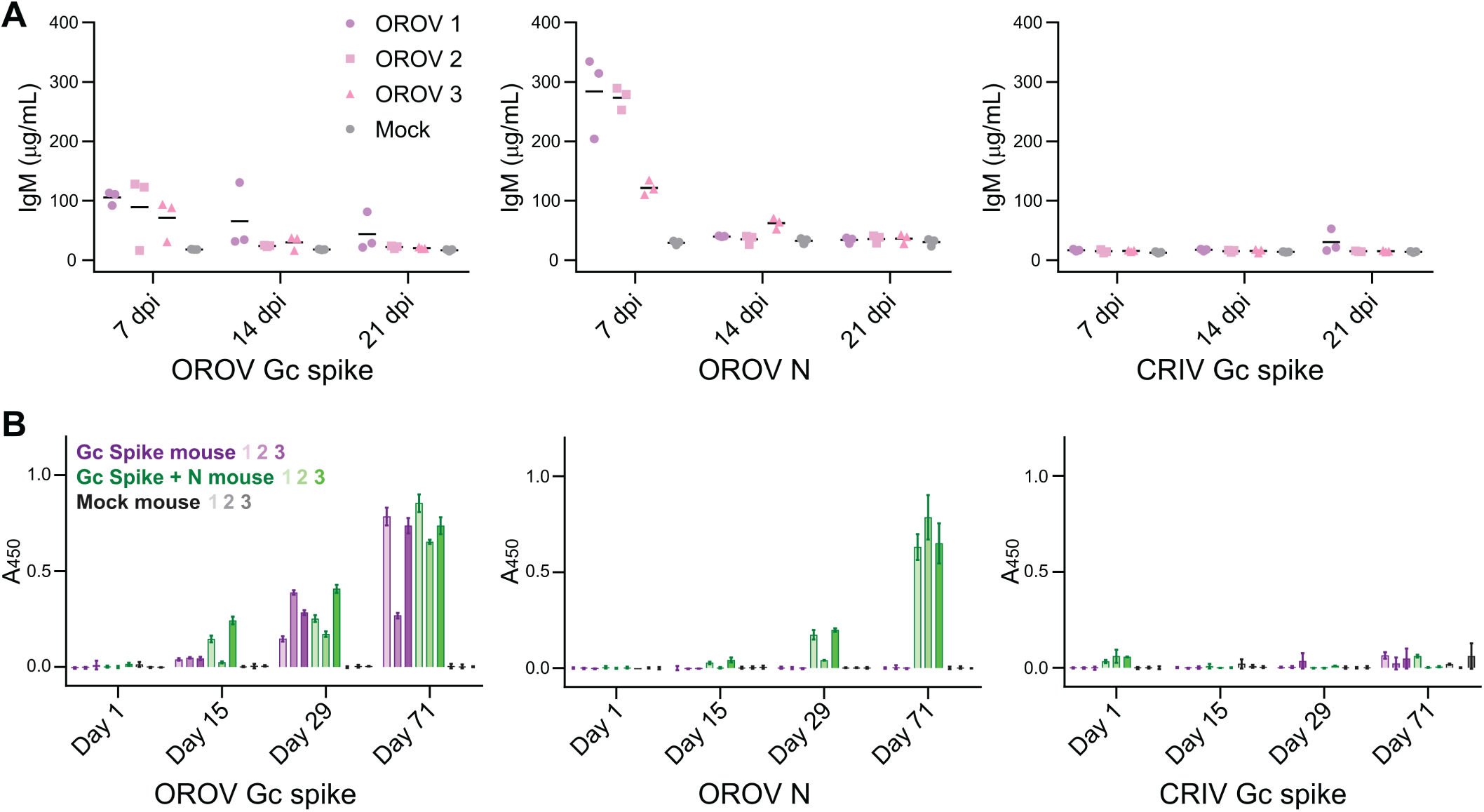
Purified OROV antigens are recognised by antibodies raised in response to OROV infection, and purified antigens raise an immune response in mice. (**A**) Blood was harvested at indicated days post-infection (dpi) and an indirect ELISA to detect IgM was performed using indicated antigens. Values are show for three independent ELISA experiments using serum from three infected mice or one mock-infected control mouse. (**B**) Pre-boost serum was collected from mice immunised at day 1 and the boosted at days 15 and 29 with purified OROV antigens Gc spike alone (purple), OROV Gc spike plus N (green), or mock-immunised (3 mice per treatment). Serum was also collected 6 weeks after the final boost. The presence of antibodies that recognise OROV Gc spike, OROV N, or CRIV Gc spike (negative control) was tested by indirect ELISA. Mean ± SD of three independent measurements for each mouse is shown.

**Figure S2.**
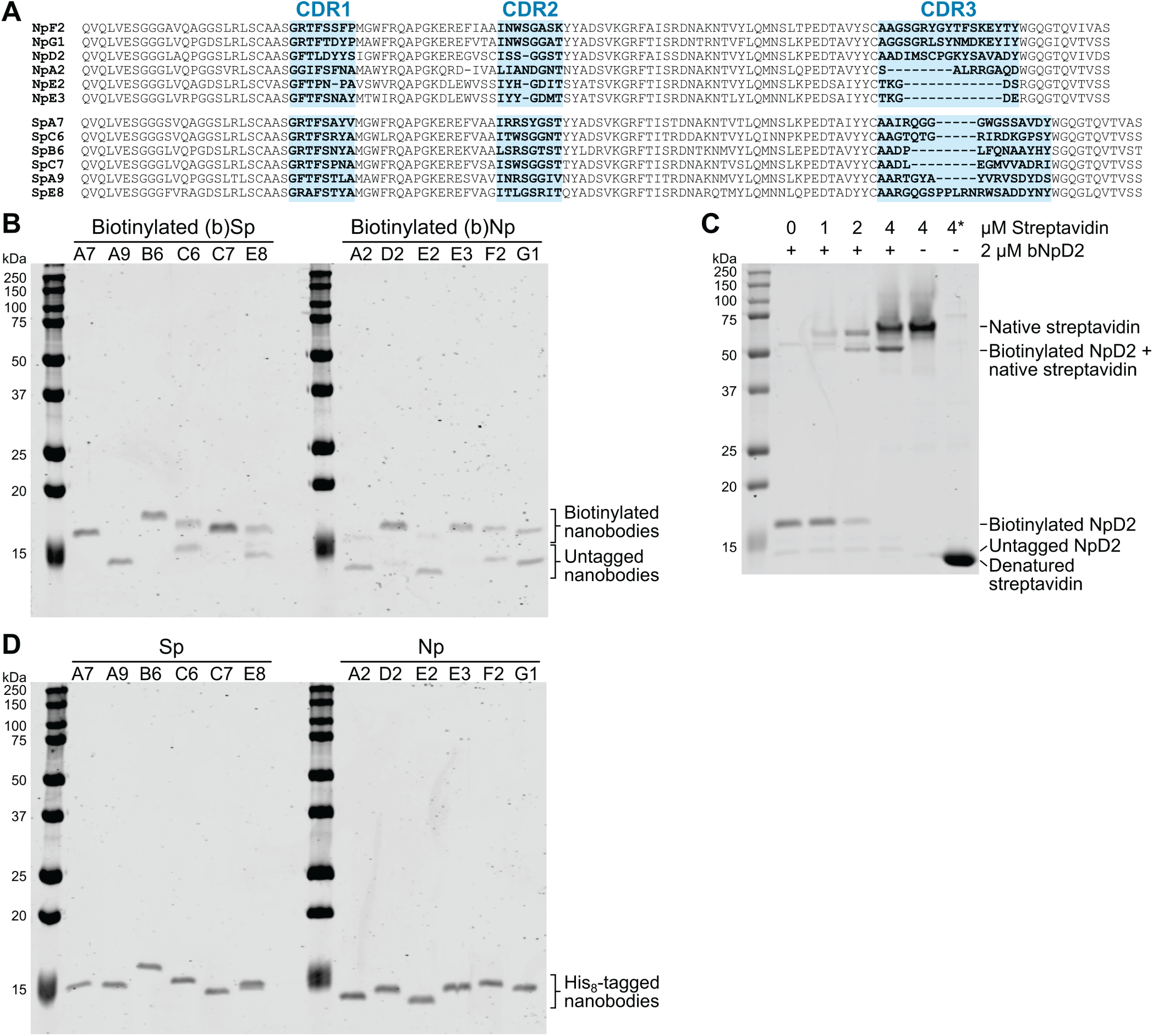
Generation and purification of nanobodies to detect OROV Gc spike and N. (**A**) Amino acid sequence alignment of selected nanobodies that recognise OROV N (NpXX) or Gc spike (SpXX). Complementarity Determining Regions (CDRs) as defined by IGMT [32] are highlighted. (**B**) Coomassie-stained SDS-PAGE of nanobodies that had been biotinylated *in vitro*. Upper bands represent the Avi-tagged, biotinylated nanobodies while lower bands represent nanobodies where the tag had been lost, presumably by proteolysis during the purification procedure. (**C**) Electrophoretic mobility shift assay to confirm biotinylation. After boiling of the nanobody in SDS-PAGE loading buffer, streptavidin was added at a 2:1, 1:1 or 1:2 molar ratio (nanobody:streptavidin) and samples were subjected to SDS-PAGE. Appearance of a high apparent molecular weight band, and disappearance of the low molecular weight band, confirms biotinylation of the nanobody. Asterisk (*) denotes streptavidin that was boiled before SDS-PAGE, rather than being added to the sample buffer after boiling.

**Figure S3.**
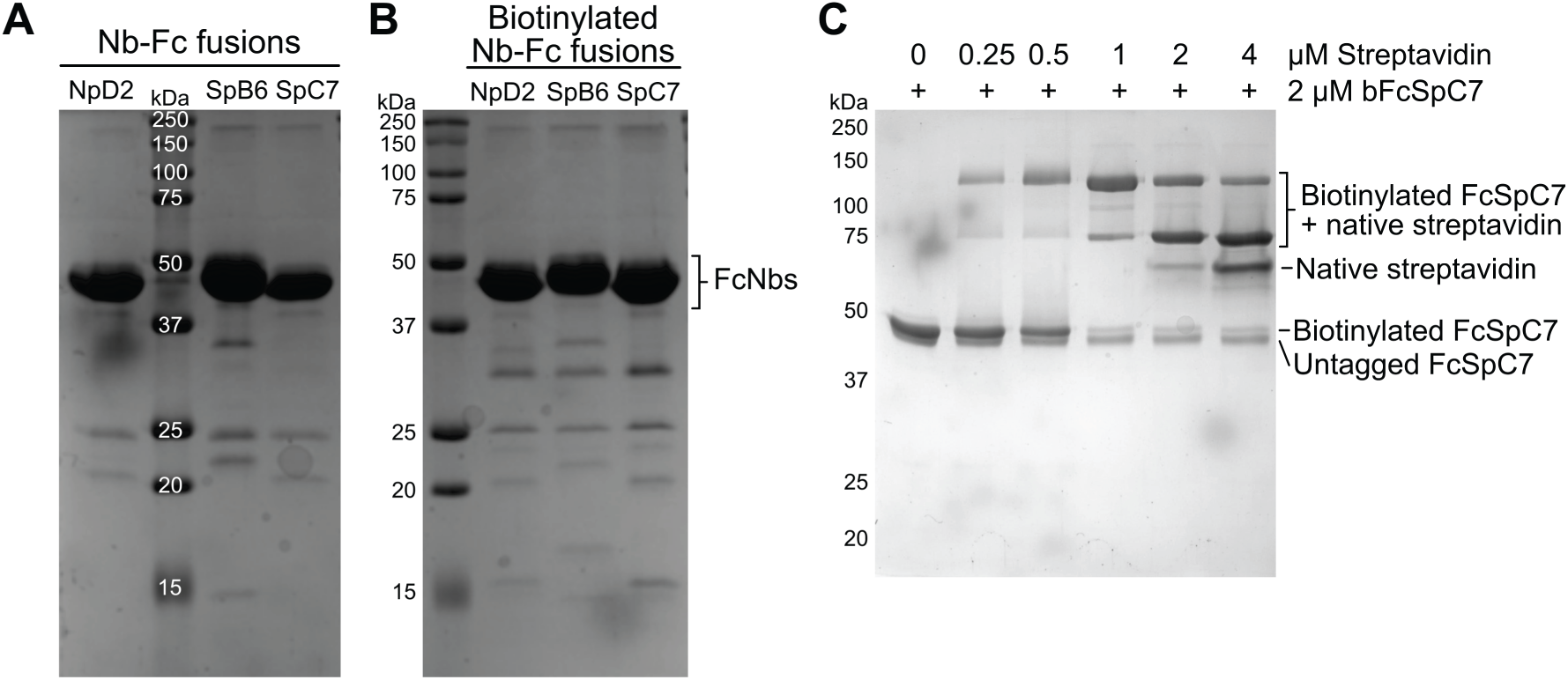
Purification of nanobody-Fc fusions. (**A, B**) Coomassie-stained SDS-PAGE of (**A**) untagged nanobody-Fc fusions, and (**B**) biotinylated nanobody-Fc fusions. (**C**) Electrophoretic mobility shift assay to confirm biotinylation. After boiling of the nanobody-Fc fusion in SDS-PAGE loading buffer, streptavidin was added at a 8:1, 4:1, 2:1, 1:1 or 1:2 molar ratio (nanobody-Fc fusion:streptavidin) and samples were subjected to SDS-PAGE. Appearance of a high apparent molecular weight bands, and reduction of the lower molecular weight band, confirms biotinylation of the nanobody-Fc fusion.

**Figure S4.**
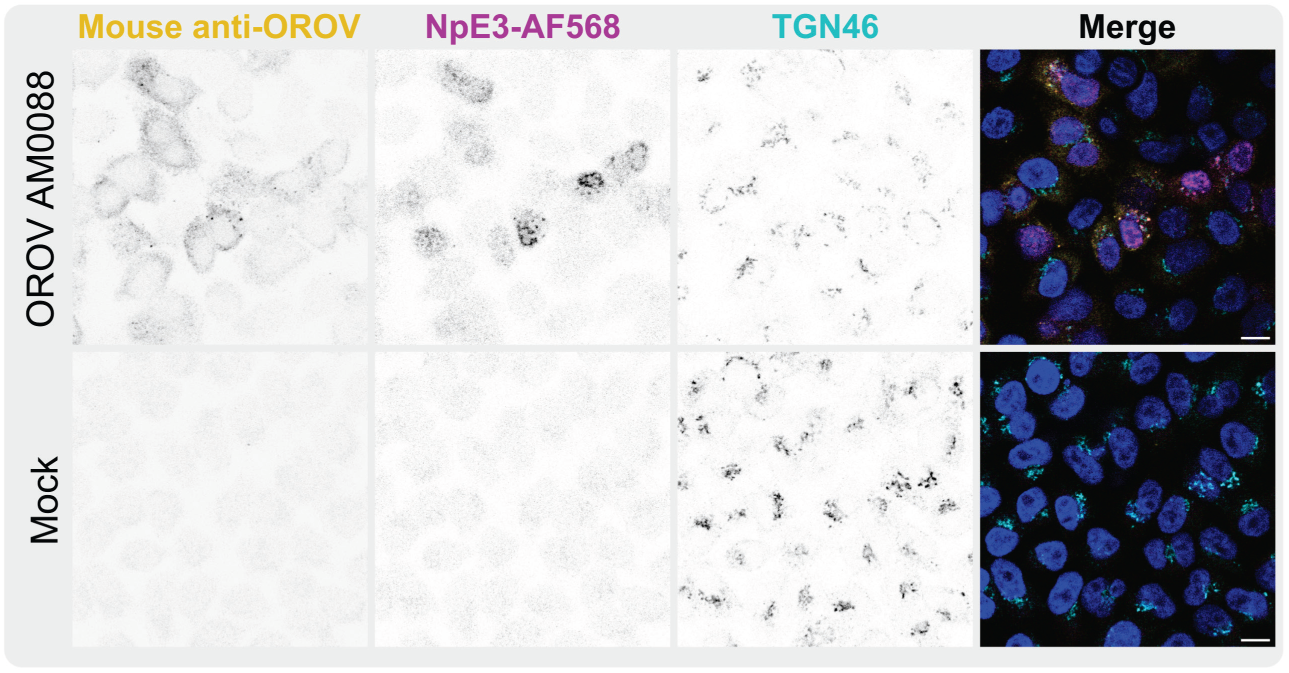
Nanobodies detect infection of HeLa cells with new OROV isolate AM0088. HeLa cells were infected with OROV AM0088 (MOI 0.5). Cells were probed with AlexaFluor (AF)568 conjugated OROV nanobody NpE3, with a polyclonal antibody against OROV and with an antibody against TGN46, plus appropriate secondary antibodies. Nuclei are stained with DAPI (blue). Scale bar = 10 µm.

## References

1. Travassos da Rosa JF, de Souza WM, Pinheiro F de P, Figueiredo ML, Cardoso JF, Acrani GO, et al. Oropouche Virus: Clinical, Epidemiological, and Molecular Aspects of a Neglected Orthobunyavirus. Am J Trop Med Hyg. 2017;96: 1019–1030. doi:10.4269/ajtmh.16-0672

2. Sakkas H, Bozidis P, Franks A, Papadopoulou C. Oropouche Fever: A Review. Viruses. 2018;10: E175. doi:10.3390/v10040175

3. Anderson CR, Spence L, Downs WG, Aitken TH. Oropouche virus: a new human disease agent from Trinidad, West Indies. Am J Trop Med Hyg. 1961;10: 574–578. doi:10.4269/ajtmh.1961.10.574

4. Mourão MPG, Bastos MS, Gimaqu JBL, Mota BR, Souza GS, Grimmer GHN, et al. Oropouche fever outbreak, Manaus, Brazil, 2007-2008. Emerg Infect Dis. 2009;15: 2063–2064. doi:10.3201/eid1512.090917

5. Bastos M de S, Figueiredo LTM, Naveca FG, Monte RL, Lessa N, Pinto de Figueiredo RM, et al. Identification of Oropouche Orthobunyavirus in the cerebrospinal fluid of three patients in the Amazonas, Brazil. Am J Trop Med Hyg. 2012;86: 732–735. doi:10.4269/ajtmh.2012.11-0485

6. Vernal S, Martini CCR, da Fonseca BAL. Oropouche Virus-Associated Aseptic Meningoencephalitis, Southeastern Brazil. Emerg Infect Dis. 2019;25: 380–382. doi:10.3201/eid2502.181189

7. Wesselmann KM, Postigo-Hidalgo I, Pezzi L, de Oliveira-Filho EF, Fischer C, de Lamballerie X, et al. Emergence of Oropouche fever in Latin America: a narrative review. Lancet Infect Dis. 2024;24: e439–e452. doi:10.1016/S1473-3099(23)00740-5

8. Files MA, Hansen CA, Herrera VC, Schindewolf C, Barrett ADT, Beasley DWC, et al. Baseline mapping of Oropouche virology, epidemiology, therapeutics, and vaccine research and development. NPJ Vaccines. 2022;7: 38. doi:10.1038/s41541-022-00456-2

9. Durango-Chavez HV, Toro-Huamanchumo CJ, Silva-Caso W, Martins-Luna J, Aguilar-Luis MA, Del Valle-Mendoza J, et al. Oropouche virus infection in patients with acute febrile syndrome: Is a predictive model based solely on signs and symptoms useful? PLoS One. 2022;17: e0270294. doi:10.1371/journal.pone.0270294

10. World Health Organization. Oropouche virus disease - Region of the Americas. 23 Aug 2024 [cited 17 Sep 2024]. Available: https://www.who.int/emergencies/disease-outbreak-news/item/2024-DON530

11. Gräf T, Delatorre E, do Nascimento Ferreira C, Rossi A, Santos HGG, Pizzato BR, et al. Expansion of Oropouche virus in non-endemic Brazilian regions: analysis of genomic characterisation and ecological drivers. Lancet Infect Dis. 2024; S1473–3099(24)00687-X. doi:10.1016/S1473-3099(24)00687-X

12. Scachetti GC, Forato J, Claro IM, Hua X, Salgado BB, Vieira A, et al. Re-emergence of Oropouche virus between 2023 and 2024 in Brazil: an observational epidemiological study. Lancet Infect Dis. 2025;25: 166–175. doi:10.1016/S1473-3099(24)00619-4

13. Naveca FG, Almeida TAP de, Souza V, Nascimento V, Silva D, Nascimento F, et al. Human outbreaks of a novel reassortant Oropouche virus in the Brazilian Amazon region. Nat Med. 2024;30: 3509–3521. doi:10.1038/s41591-024-03300-3

14. Martins-Filho PR, Carvalho TA, Dos Santos CA. Oropouche fever: reports of vertical transmission and deaths in Brazil. Lancet Infect Dis. 2024; S1473–3099(24)00557–7. doi:10.1016/S1473-3099(24)00557-7

15. Garcia Filho C, Lima Neto AS, Maia AMPC, da Silva LOR, Cavalcante R da C, Monteiro H da S, et al. A Case of Vertical Transmission of Oropouche Virus in Brazil. N Engl J Med. 2024;391: 2055–2057. doi:10.1056/NEJMc2412812

16. Bandeira AC, Pereira FM, Leal A, Santos SPO, Barbosa AC, Souza MSPL, et al. Fatal Oropouche Virus Infections in Nonendemic Region, Brazil, 2024. Emerg Infect Dis. 2024;30. doi:10.3201/eid3011.241132

17. Weidmann M, Rudaz V, Nunes MRT, Vasconcelos PFC, Hufert FT. Rapid detection of human pathogenic orthobunyaviruses. J Clin Microbiol. 2003;41: 3299–3305. doi:10.1128/JCM.41.7.3299-3305.2003

18. Wise EL, Márquez S, Mellors J, Paz V, Atkinson B, Gutierrez B, et al. Oropouche virus cases identified in Ecuador using an optimised qRT-PCR informed by metagenomic sequencing. PLoS Negl Trop Dis. 2020;14: e0007897. doi:10.1371/journal.pntd.0007897

19. Pan American Health Organization (PAHO). Recommendations for the Detection and Surveillance of Oropouche in possible cases of vertical infection, congenital malformation, or fetal death. 17 Jul 2024 [cited 24 Feb 2025]. Available: https://www.paho.org/en/documents/recommendations-detection-and-surveillance-oropouche-possible-cases-vertical-infection

20. Naveca FG, Nascimento VA do, Souza VC de, Nunes BTD, Rodrigues DSG, Vasconcelos PF da C. Multiplexed reverse transcription real-time polymerase chain reaction for simultaneous detection of Mayaro, Oropouche, and Oropouche-like viruses. Mem Inst Oswaldo Cruz. 2017;112: 510–513. doi:10.1590/0074-02760160062

21. Alva-Urcia C, Aguilar-Luis MA, Palomares-Reyes C, Silva-Caso W, Suarez-Ognio L, Weilg P, et al. Emerging and reemerging arboviruses: A new threat in Eastern Peru. PLoS One. 2017;12: e0187897. doi:10.1371/journal.pone.0187897

22. Martins-Luna J, Del Valle-Mendoza J, Silva-Caso W, Sandoval I, Del Valle LJ, Palomares-Reyes C, et al. Oropouche infection a neglected arbovirus in patients with acute febrile illness from the Peruvian coast. BMC Res Notes. 2020;13: 67. doi:10.1186/s13104-020-4937-1

23. Andreolla AP, Borges AA, Nagashima S, Vaz de Paula CB, de Noronha L, Zanchin NIT, et al. Development of monoclonal antibodies against oropouche virus and its applicability to immunohistochemical diagnosis. Virol J. 2024;21: 81. doi:10.1186/s12985-024-02323-z

24. Harmsen MM, De Haard HJ. Properties, production, and applications of camelid single-domain antibody fragments. Appl Microbiol Biotechnol. 2007;77: 13–22. doi:10.1007/s00253-007-1142-2

25. Acrani GO, Tilston-Lunel NL, Spiegel M, Weidmann M, Dilcher M, Andrade da Silva DE, et al. Establishment of a minigenome system for Oropouche virus reveals the S genome segment to be significantly longer than reported previously. J Gen Virol. 2015;96: 513–523. doi:10.1099/jgv.0.000005

26. Rodriguez C, Gricourt G, Ndebi M, Demontant V, Poiteau L, Burrel S, et al. Fatal Encephalitis Caused by Cristoli Virus, an Emerging Orthobunyavirus, France. Emerg Infect Dis. 2020;26: 1287–1290. doi:10.3201/eid2606.191431

27. Teo H, Perisic O, González B, Williams RL. ESCRT-II, an endosome-associated complex required for protein sorting: crystal structure and interactions with ESCRT-III and membranes. Dev Cell. 2004;7: 559–569. doi:10.1016/j.devcel.2004.09.003

28. Li Z, Michael IP, Zhou D, Nagy A, Rini JM. Simple piggyBac transposon-based mammalian cell expression system for inducible protein production. Proc Natl Acad Sci U S A. 2013;110: 5004–5009. doi:10.1073/pnas.1218620110

29. Aricescu AR, Lu W, Jones EY. A time- and cost-efficient system for high-level protein production in mammalian cells. Acta Crystallogr D Biol Crystallogr. 2006;62: 1243–1250. doi:10.1107/S0907444906029799

30. Bushell KM, Söllner C, Schuster-Boeckler B, Bateman A, Wright GJ. Large-scale screening for novel low-affinity extracellular protein interactions. Genome Res. 2008;18: 622–630. doi:10.1101/gr.7187808

31. Eyssen LE-A, Ramadurai S, Abdelkarim S, Buckle I, Cornish K, Lin H, et al. From Llama to Nanobody: A Streamlined Workflow for the Generation of Functionalised VHHs. Bio Protoc. 2024;14: e4962. doi:10.21769/BioProtoc.4962

32. Giudicelli V, Brochet X, Lefranc M-P. IMGT/V-QUEST: IMGT Standardized Analysis of the Immunoglobulin (IG) and T Cell Receptor (TR) Nucleotide Sequences. Cold Spring Harb Protoc. 2011;2011: pdb.prot5633. doi:10.1101/pdb.prot5633

33. Fairhead M, Howarth M. Site-specific biotinylation of purified proteins using BirA. Methods Mol Biol. 2015;1266: 171–184. doi:10.1007/978-1-4939-2272-7_12

34. Aquino VH, Moreli ML, Moraes Figueiredo LT. Analysis of oropouche virus L protein amino acid sequence showed the presence of an additional conserved region that could harbour an important role for the polymerase activity. Arch Virol. 2003;148: 19–28. doi:10.1007/s00705-002-0913-4

35. Girt GC, Lakshminarayanan A, Huo J, Dormon J, Norman C, Afrough B, et al. The use of nanobodies in a sensitive ELISA test for SARS-CoV-2 Spike 1 protein. R Soc Open Sci. 2021;8: 211016. doi:10.1098/rsos.211016

36. Concha JO, Gutierrez K, Barbosa N, Rodrigues RL, de Carvalho AN, Tavares LA, et al. Rab27a GTPase and its effector Myosin Va are host factors required for efficient Oropouche virus cell egress. PLoS Pathog. 2024;20: e1012504. doi:10.1371/journal.ppat.1012504

37. Hughes HR, Adkins S, Alkhovskiy S, Beer M, Blair C, Calisher CH, et al. ICTV Virus Taxonomy Profile: Peribunyaviridae. J Gen Virol. 2020;101: 1–2. doi:10.1099/jgv.0.001365

38. Bowden TA, Bitto D, McLees A, Yeromonahos C, Elliott RM, Huiskonen JT. Orthobunyavirus ultrastructure and the curious tripodal glycoprotein spike. PLoS Pathog. 2013;9: e1003374. doi:10.1371/journal.ppat.1003374

39. Hellert J, Aebischer A, Wernike K, Haouz A, Brocchi E, Reiche S, et al. Orthobunyavirus spike architecture and recognition by neutralizing antibodies. Nat Commun. 2019;10: 879. doi:10.1038/s41467-019-08832-8

40. Hover S, Charlton FW, Hellert J, Swanson JJ, Mankouri J, Barr JN, et al. Organisation of the orthobunyavirus tripodal spike and the structural changes induced by low pH and K+ during entry. Nat Commun. 2023;14: 5885. doi:10.1038/s41467-023-41205-w

41. Saeed MF, Nunes M, Vasconcelos PF, Travassos Da Rosa AP, Watts DM, Russell K, et al. Diagnosis of Oropouche virus infection using a recombinant nucleocapsid protein-based enzyme immunoassay. J Clin Microbiol. 2001;39: 2445–2452. doi:10.1128/JCM.39.7.2445-2452.2001

42. Barbosa NS, Concha JO, daSilva LLP, Crump CM, Graham SC. Oropouche Virus Glycoprotein Topology and Cellular Requirements for Glycoprotein Secretion. J Virol. 2023;97: e0133122. doi:10.1128/jvi.01331-22

43. Murillo JL, Cabral AD, Uehara M, da Silva VM, Dos Santos JV, Muniz JRC, et al. Nucleoprotein from the unique human infecting Orthobunyavirus of Simbu serogroup (Oropouche virus) forms higher order oligomers in complex with nucleic acids in vitro. Amino Acids. 2018;50: 711–721. doi:10.1007/s00726-018-2560-4

44. Richard G, Meyers AJ, McLean MD, Arbabi-Ghahroudi M, MacKenzie R, Hall JC. In vivo neutralization of α-cobratoxin with high-affinity llama single-domain antibodies (VHHs) and a VHH-Fc antibody. PLoS One. 2013;8: e69495. doi:10.1371/journal.pone.0069495

45. Barbosa NS, Mendonça LR, Dias MVS, Pontelli MC, da Silva EZM, Criado MF, et al. ESCRT machinery components are required for Orthobunyavirus particle production in Golgi compartments. PLoS Pathog. 2018;14: e1007047. doi:10.1371/journal.ppat.1007047

46. Watts DM, Russell KL, Wooster MT, Sharp TW, Morrison AC, Kochel TJ, et al. Etiologies of Acute Undifferentiated Febrile Illnesses in and near Iquitos from 1993 to 1999 in the Amazon River Basin of Peru. Am J Trop Med Hyg. 2022;107: 1114–1128. doi:10.4269/ajtmh.22-0259

47. Vasconcelos HB, Azevedo RSS, Casseb SM, Nunes-Neto JP, Chiang JO, Cantuária PC, et al. Oropouche fever epidemic in Northern Brazil: epidemiology and molecular characterization of isolates. J Clin Virol. 2009;44: 129–133. doi:10.1016/j.jcv.2008.11.006

48. das Neves Martins FE, Chiang JO, Nunes BTD, Ribeiro B de FR, Martins LC, Casseb LMN, et al. Newborns with microcephaly in Brazil and potential vertical transmission of Oropouche virus: a case series. Lancet Infect Dis. 2025;25: 155–165. doi:10.1016/S1473-3099(24)00617-0

49. Ladner JT, Savji N, Lofts L, Travassos da Rosa A, Wiley MR, Gestole MC, et al. Genomic and phylogenetic characterization of viruses included in the Manzanilla and Oropouche species complexes of the genus Orthobunyavirus, family Bunyaviridae. J Gen Virol. 2014;95: 1055–1066. doi:10.1099/vir.0.061309-0

50. Stubbs SH, Cornejo Pontelli M, Mishra N, Zhou C, de Paula Souza J, Mendes Viana RM, et al. Vesicular Stomatitis Virus Chimeras Expressing the Oropouche Virus Glycoproteins Elicit Protective Immune Responses in Mice. mBio. 2021;12: e0046321. doi:10.1128/mBio.00463-21

51. Wernike K, Aebischer A, Roman-Sosa G, Beer M. The N-terminal domain of Schmallenberg virus envelope protein Gc is highly immunogenic and can provide protection from infection. Sci Rep. 2017;7: 42500. doi:10.1038/srep42500

52. Schwarz MM, Price DA, Ganaie SS, Feng A, Mishra N, Hoehl RM, et al. Oropouche orthobunyavirus infection is mediated by the cellular host factor Lrp1. Proc Natl Acad Sci U S A. 2022;119: e2204706119. doi:10.1073/pnas.2204706119

53. Loreau V, Rees R, Chan EH, Taxer W, Gregor K, Mußil B, et al. A nanobody toolbox to investigate localisation and dynamics of Drosophila titins and other key sarcomeric proteins. eLife. 2023;12: e79343. doi:10.7554/eLife.79343

